# Loneliness, Functional Rurality, and Wearable-Measured Physical Activity and Sleep in the All of Us Research Program

**DOI:** 10.64898/2026.04.08.26350412

**Authors:** Shuai Yang, Jingyue Wu, Yann C. Klimentidis, David A. Sbarra

## Abstract

Loneliness—the perceived discrepancy between desired and actual social connection—is a common and aversive psychological state associated with a range of adverse health outcomes. Several theoretical models suggest that these associations may operate partly through health behaviors. In this preregistered study, we used data from the All of Us Research Program to evaluate associations of loneliness and functional rurality (FR), a study-specific contextual index of reduced neighborhood accessibility, with Fitbit-derived physical activity and sleep outcomes. Final samples included 16,912 participants for physical activity analyses and 13,937 for sleep analyses. In adjusted models, higher FR was associated with greater loneliness (β = 0.061, 95% CI [0.045, 0.077], p = 9.63 × 10^−14^). FR and loneliness were independently associated with fewer daily steps and lower moderate-to-vigorous physical activity. Loneliness was also associated with shorter sleep duration, greater sleep duration variability, higher odds of short sleep, and higher odds of low sleep efficiency. FR was not associated with sleep duration or sleep duration variability but showed a small positive association with mean sleep efficiency and lower odds of low sleep efficiency. Interaction analyses provided little evidence that FR modified the associations of loneliness with most outcomes, although the FR × loneliness interaction was significant for sleep duration variability, indicating that loneliness was more strongly associated with irregular sleep duration in higher-FR contexts. Sensitivity analyses using stricter valid-day thresholds, winsorization, quartile-based exposure coding, and a backward 30-day window yielded directionally similar findings. These results suggest that FR and loneliness are independently associated with lower physical activity, whereas loneliness shows a more consistent relationship with adverse sleep patterns.

---

Loneliness is a common and aversive psychological state that is associated with poor mental and physical health, elevated risk of morbidity from a range of disease outcomes, and premature mortality.^1,2^ The overall strength and magnitude of the association between loneliness and health has led the U.S. Surgeon General to identify loneliness as a public health crisis.^3^ Although loneliness is often framed as an individual psychological experience, it is also embedded in broader social and environmental contexts that shape opportunities for social connection, mobility, and daily routine.^4,5^

Many models linking loneliness to distal health do so via everyday health behaviors, and this work often cites disturbed sleep and limited physical activity as potential mechanistic processes.^1,4^ Loneliness is linked with poorer sleep, particularly self-reported sleep disturbance,^5^ and evolutionary accounts of loneliness describe disturbed sleep as an adaptive social-vigilance system for assessing one’s standing on the social periphery.^6^ Prior empirical work, for example, finds that objectively measured loneliness prospectively predicts greater sleep fragmentation and lower sleep efficiency, suggesting that perceived isolation triggers a state of heightened physiological arousal that disrupts restorative sleep.^7^ Associations with physical activity are less consistent, but prior reviews suggest that social resources and perceived isolation may influence engagement in regular activity, especially in older adults.^8^ For example, Schrempft et al. demonstrated that socially isolated individuals engaged in less objectively measured moderate-to-vigorous physical activity and more sedentary behavior, consistent with the idea that social engagement may help support daily movement.^9^

The broader built environment may also shape loneliness and health behavior. Reviews of built-environment research indicate that neighborhood accessibility, connected infrastructure, and proximity to destinations are important correlates of walking and routine movement.^10^ More recently, work on loneliness and place has suggested that restrictive built environments may contribute to loneliness by limiting mobility, social contact, access to services, and use of shared spaces; in this sense, some environments may be considered “lonelygenic” because they constrain opportunities for everyday social connection.^11^ In the present study, we define functional rurality (FR) as a study-specific contextual index of reduced neighborhood access to walkable shops, public transit, sidewalks, bicycle facilities, and low-cost recreation. We use FR to capture a more rural-like accessibility context rather than geographic rural residence per se.

FR may act as a structural moderator in behavioral pathways linking loneliness to health because neighborhood accessibility helps shape opportunities for incidental social contact.^12,13^ In highly walkable, resource-dense neighborhoods (low FR), individuals may have more opportunities for routine encounters and the development of “weak ties” that support social embeddedness.^12,13^ These everyday interactions may help buffer some of the adverse psychological and behavioral correlates of loneliness.^12,13^ By contrast, high-FR environments, characterized by poorer accessibility, may limit these structural opportunities for connection and thereby strengthen the association between loneliness and behavioral withdrawal. From this perspective, the present study examines how structural and functional elements of social connectedness may intersect in shaping health-relevant behaviors.

This framing is conceptually grounded in neighborhood environment research and emerges from recent methodological work validating the Social Determinants of Health (SDoH) surveys within the All of Us Research Program.^14,15^ All of Us is a large-scale, nationwide precision medicine initiative aimed at building a diverse biomedical data resource. The SDoH survey includes items assessing everyday neighborhood infrastructure, and recent methodological work has used these items to operationalize the FR measure examined here.^15,16^ Its face and content validity are supported by prior neighborhood-environment research, particularly PANES (the Physical Activity Neighborhood Environment Scale), which was developed as a brief measure of walkability- and activity-relevant neighborhood features and demonstrated acceptable reliability for research and surveillance use.^14,17,18^ This framing of FR also has precedent in scholarship on rural accessibility and service context, including work that emphasizes access to services, mobility, and spatial structure as important features of lived rural environments.^17,18^ We therefore treat FR in this study as an effort to operationalize and evaluate a structural accessibility-oriented rurality construct in a large and diverse U.S. cohort.

Beyond the assessment of FR, the All of Us Research Program provides a valuable opportunity to examine daily sleep and physical activity using wearable technology, helping to reduce shared method variance between loneliness and health behavior measures. The All of Us Research Program is well suited to this aim because it combines large-scale survey data with wearable-device data in a diverse U.S. cohort.^19^ Wearable devices enable passive measurement of free-living activity and sleep and show acceptable validity for key behavioral indicators, including step counts, activity intensity, and sleep duration.^20,21^ Integrating objective behavioral metrics with comprehensive psychosocial data provides a strong approach for testing behavioral pathways. Prior studies have linked loneliness and social isolation to objectively measured sleep and physical activity, including actigraph-assessed sleep fragmentation and sleep quality as well as accelerometer-measured physical activity and sedentary behavior.^7,9,22,23^ The All of Us Research Program extends this literature by enabling these questions to be examined in a large and diverse U.S. cohort using wearable-derived measures of free-living sleep and physical activity.

## The Present Study

Drawing on data from the SDoH survey and Fitbit daily summaries in All of Us, we examined three preregistered (OSF: https://osf.io/9857p/overview?view_only=477e2e2f76c340fe8aadbee284064f07) research questions: (1) whether higher FR is associated with greater loneliness; (2) whether FR and loneliness are independently associated with Fitbit-derived physical activity and sleep outcomes; and (3) whether FR modifies the associations between loneliness and those outcomes. Based on prior literature, we hypothesized that higher FR is associated with greater loneliness, that both FR and loneliness are associated with lower physical activity and worse sleep, and that associations of loneliness with health behaviors are stronger in higher-FR contexts.

## Methods

### Study Design and Data Source

We conducted a cross-sectional analysis within the All of Us Controlled Tier Dataset v8. The All of Us Research Program is a large U.S. cohort that integrates survey, electronic health record, physical measurement, biospecimen, and digital health data within a secure cloud-based research workbench.^19^ Exposures were derived from the SDoH survey, which launched on November 1, 2021,^15^ and behavioral outcomes were derived from Fitbit daily activity and sleep summary data. We aligned exposures and outcomes using a person-specific index date (*t0*) defined from the SDoH survey and evaluated Fitbit outcomes within a prespecified 30-day observation window relative to that date. Although the study was preregistered, all key design and analytic decisions are described here to make the methods section fully self-contained.

### Participant Selection

Participants were eligible if they were aged 18 years or older and had at least one SDoH survey completed on or after November 1, 2021. To be eligible for exposure assessment, the same SDoH survey instance had to contain sufficient information to score both FR and loneliness. FR required at least 3 of 5 non-missing items, and loneliness required at least 4 of 8 non-missing items. For each participant, the earliest SDoH survey instance meeting both scoring rules was designated as *t0*. To maintain a consistent analytic base across the preregistered research questions, eligibility required that the same SDoH survey instance contain sufficient data to score both FR and loneliness, even for models in which only one of the two exposures was entered.

Fitbit outcomes were assessed during the forward 30-day window from *t0* through *t0* + 29 days. Participants did not need Fitbit data on the exact *t0* date or on all 30 days; inclusion required only sufficient valid days or nights within that person-specific window. Participants were included in the physical activity analytic sample if they had at least 7 valid physical activity days and complete covariate data. Participants were included in the sleep analytic sample if they had at least 7 valid sleep nights and complete covariate data. Cohort construction is shown in Figure 1.

**Figure 1.**
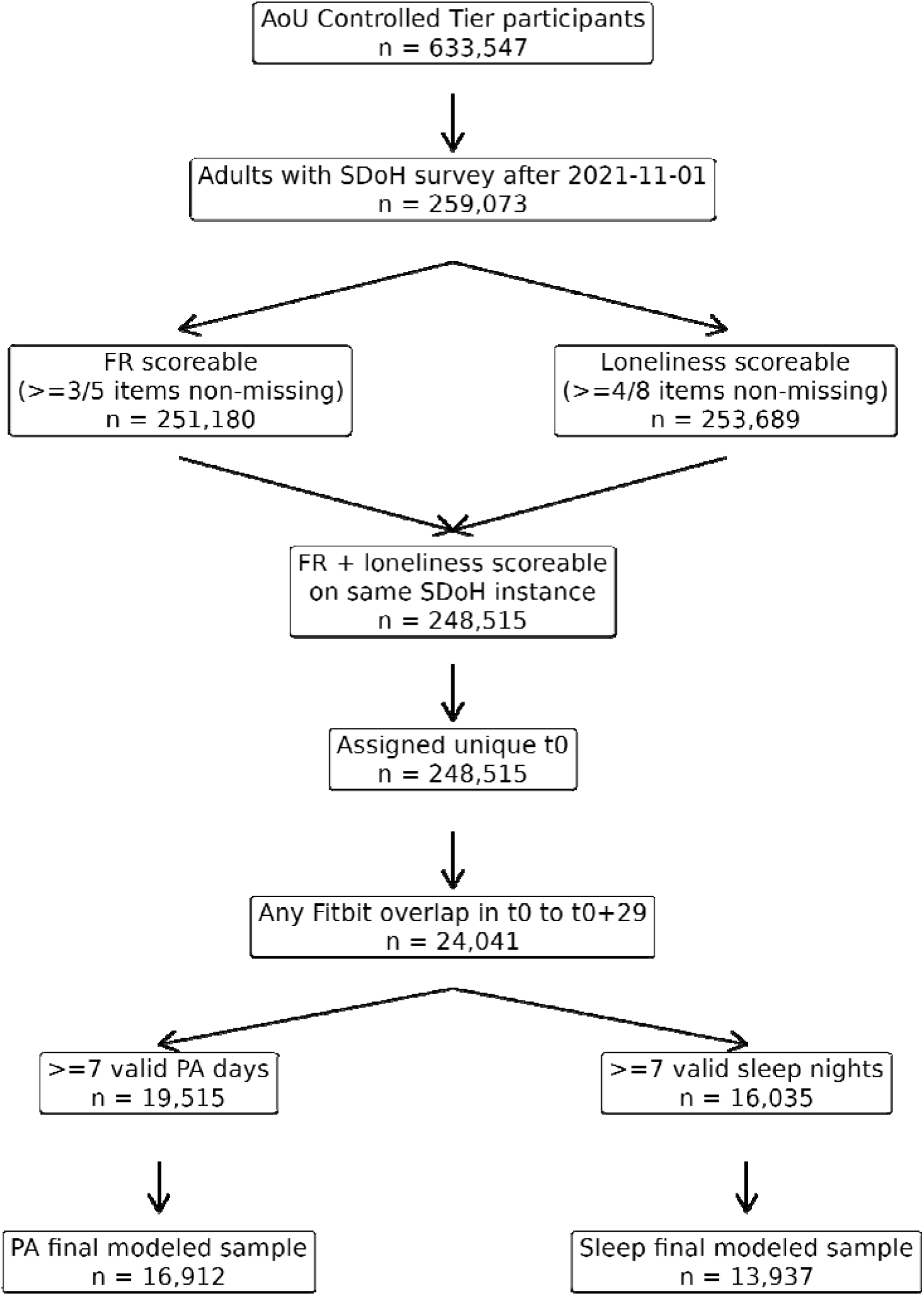
Cohort construction for the functional rurality–loneliness–Fitbit analytic sample.

### Measures

#### Functional Rurality

FR was derived from five neighborhood accessibility and built-environment items from the SDoH survey: availability of walkable shops, public transit within a 10–15 minute walk, sidewalks, bicycle facilities, and free or low-cost recreation facilities. Items were coded so that higher values reflected poorer access and a more rural-like accessibility context. The FR score was calculated as the mean of available items when at least 3 of 5 items were non-missing.

Because the SDoH survey combines items from multiple established sources, FR should be interpreted as a theory-informed, study-specific contextual index rather than an official validated All of Us scale.^15^

#### Loneliness

Loneliness was assessed using eight SDoH survey items corresponding to the UCLA Loneliness Scale short form. Positively worded items were reverse-coded so that higher values indicated greater loneliness. The loneliness score was calculated as the mean of available items when at least 4 of 8 items were non-missing.^24^

#### Fitbit Outcomes: Physical Activity and Sleep

Two continuous physical activity outcomes were examined: mean daily steps and mean daily moderate-to-vigorous physical activity (MVPA) minutes during the 30-day window after *t0*. A valid physical activity day was defined as a day with at least 100 recorded steps. For each participant, daily values were averaged across all valid days in the analytic window.

Sleep outcomes included mean nightly sleep duration (hours), mean sleep efficiency, sleep duration variability, short sleep, and low sleep efficiency over the same 30-day window. Sleep outcomes were derived from Fitbit daily sleep summaries and restricted to main sleep episodes.

Sleep duration was defined as nightly total sleep time from the Fitbit daily sleep summary and converted to hours. Sleep efficiency was defined as the proportion of time asleep while in bed during the main sleep episode. Sleep duration variability was defined as the within-person standard deviation of nightly sleep duration across valid nights, reflecting night-to-night instability in sleep duration. Short sleep was defined as mean sleep duration < 7 hours, consistent with adult sleep duration recommendations.^25^ Low sleep efficiency was defined as mean sleep efficiency < .85. This cutoff was selected because sleep efficiency of 85% or higher has been identified as an indicator of good sleep quality in expert-consensus recommendations, and values below 85% have commonly been used as a pragmatic indicator of poorer sleep continuity in population-based sleep research.^26^ Given the use of consumer wearable data, this threshold should be interpreted as a pragmatic descriptive cutoff rather than a clinical diagnostic threshold.

A valid sleep night required non-missing sleep duration, non-missing sleep efficiency, and positive time in bed. Fitbit outcomes were treated as wearable-based proxies of free-living behavior rather than gold-standard clinical sleep or activity measures.^20,21,27,28^

#### Covariates

Covariates included age, gender, race, education, income, smoking status, alcohol use, and season of *t0*. Age (in years) was treated as a continuous variable and defined at *t0*, given its strong associations with loneliness, physical activity, and sleep. Sex/gender was included as a categorical variable (female, male, and non-binary). Race was self-identified and categorized into mutually exclusive groups: White, Black or African American, Asian, American Indian or Alaska Native, Native Hawaiian or other Pacific Islander, more than one race, and Middle Eastern or North African. Adjusting for race allowed us to account for potential disparities in neighborhood context, loneliness, and wearable-derived behavioral outcomes. Socioeconomic factors included years of education completed, treated as a continuous variable, and annual household income, coded in ordered categories ranging from less than $25,000 to greater than $200,000, which were included because they may confound associations of FR and loneliness with physical activity and sleep through differences in resources, living conditions, and daily opportunities for health-promoting behavior. We also adjusted for smoking status (never, former or current, and unknown) and alcohol use (never, former or current, and unknown) at or before *t0*, as both behaviors are associated with sleep and physical activity patterns. Finally, season of *t0* (categorical) was included to account for potential seasonal variation in movement and sleep.

### Statistical Analysis

All data processing and statistical analyses were conducted within the All of Us Researcher Workbench using its cloud-based Jupyter environment, and all primary analyses used complete-case samples. Continuous exposures were standardized as *z* scores for model interpretation. Descriptive statistics were summarized as mean (SD) for continuous variables and *n* (%) for categorical variables. As an additional descriptive step, we examined bivariate correlations among FR, loneliness, and primary wearable outcomes.

For research question 1, we fit a multivariable linear regression model with loneliness as the outcome and FR as the exposure. For research question 2, we fit separate multivariable linear regression models for continuous outcomes and logistic regression models for binary sleep outcomes, first modeling FR and loneliness separately and then entering both exposures simultaneously in mutually adjusted models. All models adjusted for age, gender, race, education, income, smoking, alcohol use, and season of *t0*. Robust HC3 heteroskedasticity-consistent standard errors were used to reduce sensitivity to heteroskedasticity and influential observations. For research question 3, we fit interaction models including an FR × loneliness product term to test effect modification. For statistically significant interactions, we planned follow-up decomposition of the interaction using conditional effects at prespecified values of FR and graphical presentation of predicted values to aid interpretation.

Sensitivity analyses evaluated stricter wearable data thresholds (≥10 valid physical activity days or sleep nights), winsorization of continuous outcomes, quartile-based exposure coding, and a backward 30-day window before *t0*. Benjamini-Hochberg false discovery rate adjustment was applied within outcome families.^29^

## Results

### Participant Selection and Sample Characteristics

Figure 1 summarizes the cohort construction, and Table 1 presents the characteristics of the physical activity and sleep analytic samples. After primary exclusions for implausible activity and sleep values, the final modeled samples included 16,912 participants for physical activity analyses and 13,937 participants for sleep analyses. Internal consistency was acceptable to good for both study-specific scales. Cronbach’s alpha for FR was 0.800 in both the physical activity and sleep samples, whereas Cronbach’s alpha for loneliness was 0.880 in the physical activity sample and 0.879 in the sleep sample.

**Table 1.** Participant Characteristics of the Physical Activity and Sleep Analytic Samples.

| Characteristic | Physical activity sample | Sleep sample |
| --- | --- | --- |
| N | 16,912 | 13,937 |
| Age at t0, years | 54.33 (15.18) | 54.33 (15.12) |
| Functional rurality | 2.26 (0.91) | 2.26 (0.91) |
| Loneliness | 2.65 (0.59) | 2.66 (0.58) |
| Education, years | 15.91 (1.97) | 15.90 (1.97) |
| Income midpoint, USD | 97,722.48 (64,539.86) | 97,551.12 (64,494.49) |
| Valid PA days / sleep nights | 25.48 (6.54) | 23.69 (7.04) |
| Mean daily steps | 7,094.08 (3,798.50) | — |
| Mean daily MVPA minutes | 33.62 (34.76) | — |
| Mean sleep duration, hours | — | 6.46 (1.12) |
| Mean sleep efficiency | — | 0.88 (0.03) |
| Sleep duration variability, hours | — | 1.43 (0.64) |
| Female | 11,826 (69.9%) | 9,715 (69.7%) |
| Male | 4,697 (27.8%) | 3,889 (27.9%) |
| Non-binary | 389 (2.3%) | 333 (2.4%) |
| American Indian or Alaska Native | 103 (0.6%) | 76 (0.5%) |
| Asian | 641 (3.8%) | 482 (3.5%) |
| Black or African American | 1,004 (5.9%) | 736 (5.3%) |
| Middle Eastern or North African | 71 (0.4%) | 48 (0.3%) |
| More than one population | 1,033 (6.1%) | 841 (6.0%) |
| Native Hawaiian or Other Pacific Islander | <20 | <20 |
| White | 14,052 (83.1%) | 11,748 (84.3%) |
| Smoking = never | 10,781 (63.7%) | 8,837 (63.4%) |
| Smoking = former or current | 6,131 (36.3%) | 5,100 (36.6%) |
| Drinking = never | 562 (3.3%) | 454 (3.3%) |
| Drinking = former or current | 16,350 (96.7%) | 13,483 (96.7%) |
| Fall | 6,820 (40.3%) | 5,722 (41.1%) |
| Spring | 2,743 (16.2%) | 2,205 (15.8%) |
| Summer | 2,620 (15.5%) | 2,031 (14.6%) |
| Winter | 4,729 (28.0%) | 3,979 (28.5%) |
| Short sleep = 0 | — | 4,544 (32.6%) |
| Short sleep = 1 | — | 9,393 (67.4%) |
| Low sleep efficiency = 0 | — | 12,520 (89.8%) |
| Low sleep efficiency = 1 | — | 1,417 (10.2%) |

**Note.** Values are mean (SD) for continuous variables and n (%) for categorical variables. MVPA = moderate-to-vigorous physical activity.

In the physical activity analytic sample, mean age was 54.33 years (SD = 15.18), mean FR was 2.26 (SD = 0.91), and mean loneliness was 2.65 (SD = 0.59). Participants contributed a mean of 25.48 valid physical activity days (SD = 6.54), with a mean of 7,094.08 daily steps (SD = 3,798.50) and 33.62 mean daily MVPA minutes (SD = 34.76). In the sleep analytic sample, mean age was 54.33 years (SD = 15.12), mean FR was 2.26 (SD = 0.91), and mean loneliness was 2.66 (SD = 0.58). Participants contributed a mean of 23.69 valid sleep nights (SD = 7.04), with mean sleep duration of 6.46 hours (SD = 1.12), mean sleep efficiency of 0.88 (SD = 0.03), and mean sleep duration variability of 1.43 hours (SD = 0.64). Overall, 67.4% met the definition of short sleep and 10.2% met the definition of low sleep efficiency.

Correlations among the primary continuous study variables are presented visually in Supplementary Figure S1. FR was modestly positively correlated with loneliness (r = .077), modestly negatively correlated with daily steps (r = −.079) and MVPA (r = −.066), and only weakly related to mean sleep duration (r = −.008) and sleep duration variability (r = .030). Loneliness showed stronger negative correlations with daily steps (r = −.145), MVPA (r = −.097), and mean sleep duration (r = −.078), and a stronger positive correlation with sleep duration variability (r = .181).

### Association Between Functional Rurality and Loneliness

Higher functional rurality was associated with greater loneliness. In the adjusted linear regression model, each 1-SD increase in FR was associated with a 0.061-SD higher loneliness score (β = 0.061, 95% CI [0.045, 0.077], p = 9.63 × 10^−14^).

### Associations of Functional Rurality and Loneliness with Physical Activity and Sleep

Figure 2 visually summarizes the main-effect estimates from separate and mutually adjusted models; the full numerical results are provided in Supplementary Table S1. In separate models, higher FR was associated with fewer daily steps (β = −211.23, 95% CI [-268.75, −153.72], p = 6.33 × 10^−13^) and lower MVPA minutes (β = −1.78, 95% CI [-2.30, −1.25], p = 3.96 × 10^−11^). Higher loneliness was likewise associated with fewer daily steps (β = −380.06, 95% CI [-439.78, −320.35], p = 1.47 × 10^−35^) and lower MVPA minutes (β = −1.66, 95% CI [-2.19, −1.13], p = 6.19 × 10^−10^). When FR and loneliness were included simultaneously, both remained independently associated with lower physical activity. FR was associated with fewer daily steps (β = −187.49, 95% CI [-244.73, −130.24], p = 1.41 × 10^−10^) and lower MVPA minutes (β = −1.68, 95% CI [-2.20, −1.15], p = 3.84 × 10^−10^), while loneliness remained associated with fewer daily steps (β = −366.64, 95% CI [-426.27, −307.02], p = 2.56 × 10^−33^) and lower MVPA minutes (β = −1.54, 95% CI [-2.06, −1.02], p = 7.90 × 10^−9^).

**Figure 2.**
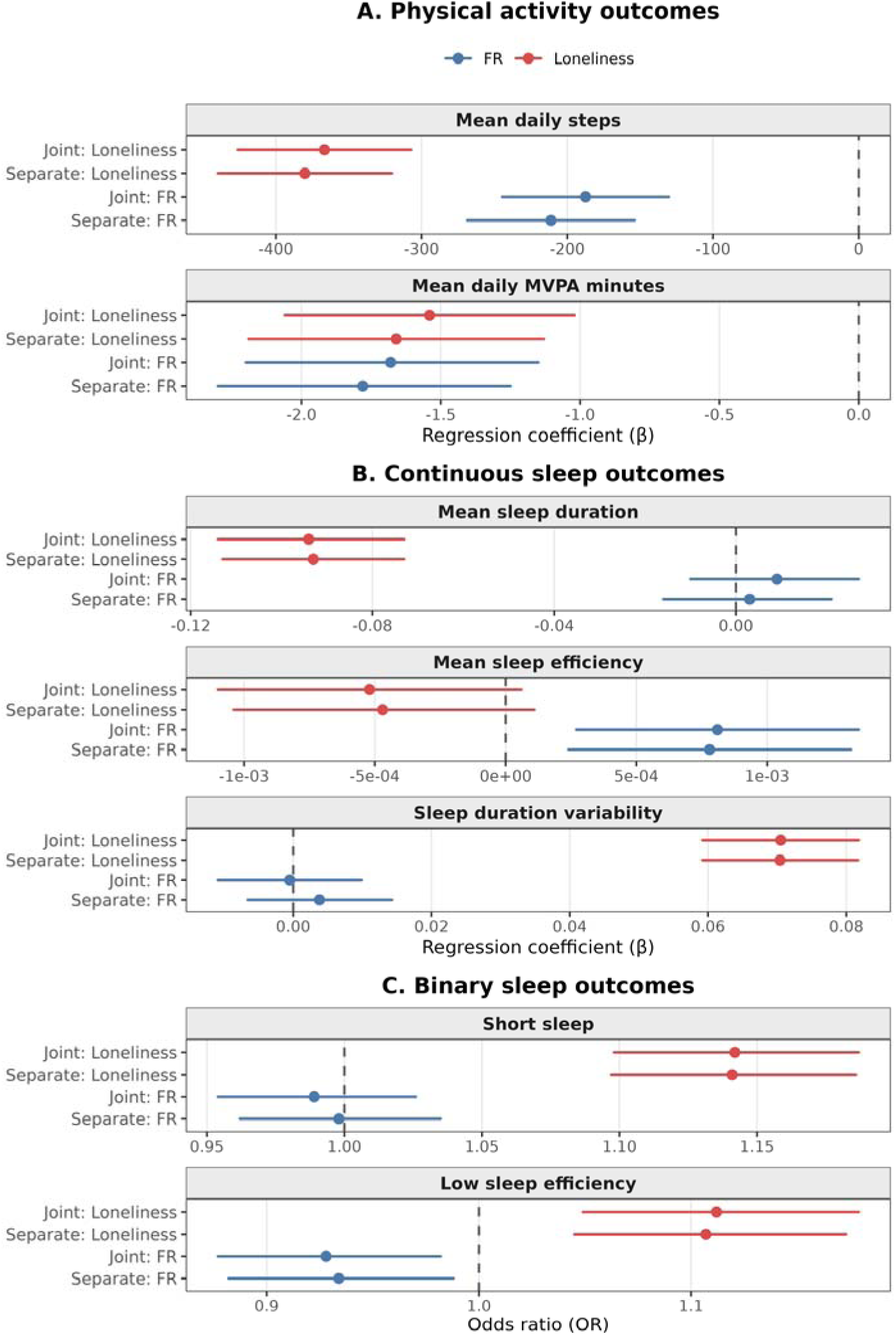
Forest plots for associations of functional rurality (FR) and loneliness with physical activity and sleep outcomes. Panel A shows results for physical activity outcomes, including mean daily steps and mean daily moderate-to-vigorous physical activity (MVPA) minutes. Panel B shows results for continuous sleep outcomes, including mean sleep duration, mean sleep efficiency, and sleep duration variability. Panel C shows results for binary sleep outcomes, including short sleep and low sleep efficiency. Separate models included FR or loneliness individually, whereas joint models included both FR and loneliness simultaneously. Points represent regression coefficients (β) for continuous outcomes or odds ratios (ORs) for binary outcomes, and horizontal lines represent 95% confidence intervals. All models were adjusted for age, gender, race, education, income, smoking, alcohol use, and season of *t0*.

For sleep outcomes, FR was not associated with mean sleep duration in separate (β = 0.003, 95% CI [-0.016, 0.021], p = .766) or mutually adjusted (β = 0.009, 95% CI [-0.010, 0.027], p = .363) models. FR was also not associated with sleep duration variability in separate (β = 0.004, 95% CI [-0.007, 0.014], p = .472) or mutually adjusted (β = −0.0005, 95% CI [-0.011, 0.010], p = .928) models. However, higher FR was associated with slightly higher mean sleep efficiency in both separate (β = 0.00078, 95% CI [0.00024, 0.00132], p = .0045) and mutually adjusted (β = 0.00081, 95% CI [0.00027, 0.00135], p = .0032) models. FR was also associated with lower odds of low sleep efficiency in both separate (OR = 0.93, 95% CI [0.88, 0.99], p = .017) and mutually adjusted (OR = 0.93, 95% CI [0.88, 0.98], p = .0099) models, but was not associated with short sleep in either separate (OR = 1.00, 95% CI [0.96, 1.03], p = .904) or mutually adjusted (OR = 0.99, 95% CI [0.95, 1.03], p = .568) models.

Higher loneliness was more consistently associated with adverse sleep. In separate models, loneliness was associated with shorter mean sleep duration (β = −0.093, 95% CI [-0.113, −0.073], p = 4.40 × 10^−19^) and greater sleep duration variability (β = 0.070, 95% CI [0.059, 0.082], p = 2.90 × 10^−34^), but not with mean sleep efficiency (β = −0.00047, 95% CI [-0.00104, 0.00011], p = .114). In logistic models, loneliness was associated with greater odds of short sleep (OR = 1.14, 95% CI [1.10, 1.19], p = 3.08 × 10^−11^) and low sleep efficiency (OR = 1.11, 95% CI [1.04, 1.17], p = 5.95 × 10^−4^). In mutually adjusted models, loneliness remained associated with shorter sleep duration (β = −0.094, 95% CI [-0.114, −0.073], p = 2.95 × 10^−19^), greater sleep duration variability (β = 0.070, 95% CI [0.059, 0.082], p = 3.43 × 10^−34^), greater odds of short sleep (OR = 1.14, 95% CI [1.10, 1.19], p = 2.72 × 10^−11^), and greater odds of low sleep efficiency (OR = 1.11, 95% CI [1.05, 1.18], p = 3.46 × 10^−4^). Loneliness was not significantly associated with mean sleep efficiency in the mutually adjusted model (β = −0.00052, 95% CI [-0.00110, 0.00006], p = .0778).

### Interaction Between Functional Rurality and Loneliness

Most FR × loneliness interaction terms were not statistically significant for physical activity or sleep outcomes (Table 2). There was no evidence of interaction for daily steps (β = −14.40, 95% CI [-70.49, 41.68], p = .615), MVPA (β = 0.283, 95% CI [-0.232, 0.798], p = .281), mean sleep duration (β = −0.0090, 95% CI [-0.0280, 0.0100], p = .354), mean sleep efficiency (β = −0.00013, 95% CI [-0.00066, 0.00039], p = .621), short sleep (OR = 1.03, 95% CI [0.993, 1.067], p = .115), or low sleep efficiency (OR = 1.01, 95% CI [0.954, 1.062], p = .807). However, the interaction term for sleep duration variability was statistically significant (β = 0.0175, 95% CI [0.0067, 0.0283], p = .00149), indicating that the positive association between loneliness and night-to-night variability in sleep duration was stronger among those in high FR settings.

**Table 2.** Interactions between functional rurality and loneliness for physical activity and sleep outcomes.

| Outcome | Interaction estimate | 95% CI | *p* |
| --- | --- | --- | --- |
| Daily steps | $\beta = -14.40$ | [-70.49, 41.68] | .615 |
| MVPA minutes | $\beta = 0.283$ | [-0.232, 0.798] | .281 |
| Sleep duration (hours) | $\beta = -0.0090$ | [-0.0280, 0.0100] | .354 |
| Sleep efficiency | $\beta = -0.00013$ | [-0.00066, 0.00039] | .621 |
| Sleep duration variability (hours) | $\beta = 0.0175$ | [0.0067, 0.0283] | .00149 |
| Short sleep | OR = 1.029 | [0.993, 1.067] | .115 |
| Low sleep efficiency | OR = 1.007 | [0.954, 1.062] | .807 |

**Note.** Interaction models included FR, loneliness, and their product term ( $FR \times loneliness$ ), with adjustment for age, gender, race, education, income, smoking, alcohol use, and season of $t_0$ . Continuous outcomes are reported as regression coefficients ( $\beta$ ), and binary outcomes are reported as odds ratios (ORs). For the significant interaction on sleep duration variability, the simple slopes for loneliness increased from $\beta = 0.0519$ at low FR to $\beta = 0.0694$ at mean FR and $\beta = 0.0869$ at high FR, indicating a stronger positive association between loneliness and sleep duration variability in higher-FR contexts.

Simple slope decomposition supported this interpretation. The association between loneliness and sleep duration variability was positive at all three tested levels of FR, but it became progressively stronger from low to high FR: at low FR (−1 SD), β = 0.0519, 95% CI [0.0359, 0.0680], p = 2.31 × 10^−10^; at mean FR, β = 0.0694, 95% CI [0.0581, 0.0807], p = 2.47 × 10^−33^; and at high FR (+1 SD), β = 0.0869, 95% CI [0.0717, 0.1020], p = 4.20 × 10^−29^. Thus, loneliness was more strongly associated with irregular sleep duration in more rural-like, lower-access contexts.

### State-Level Geographic Patterns

Figure 3 presents descriptive state-level maps of mean FR and mean loneliness in the analytic sample, and Supplementary Table S2 provides the corresponding state-level summary statistics. FR was highest in West Virginia (2.88), followed by Mississippi (2.85) and Maine (2.84), and lowest in New York (1.69), Illinois (1.76), and California (1.81). Loneliness was highest in Oklahoma (2.88), followed by Arkansas (2.84) and West Virginia (2.83), and lowest in Mississippi (2.55), Massachusetts (2.56), and California (2.57). These maps are intended to provide descriptive geographic context rather than state-level inferential comparisons.

**Figure 3.**
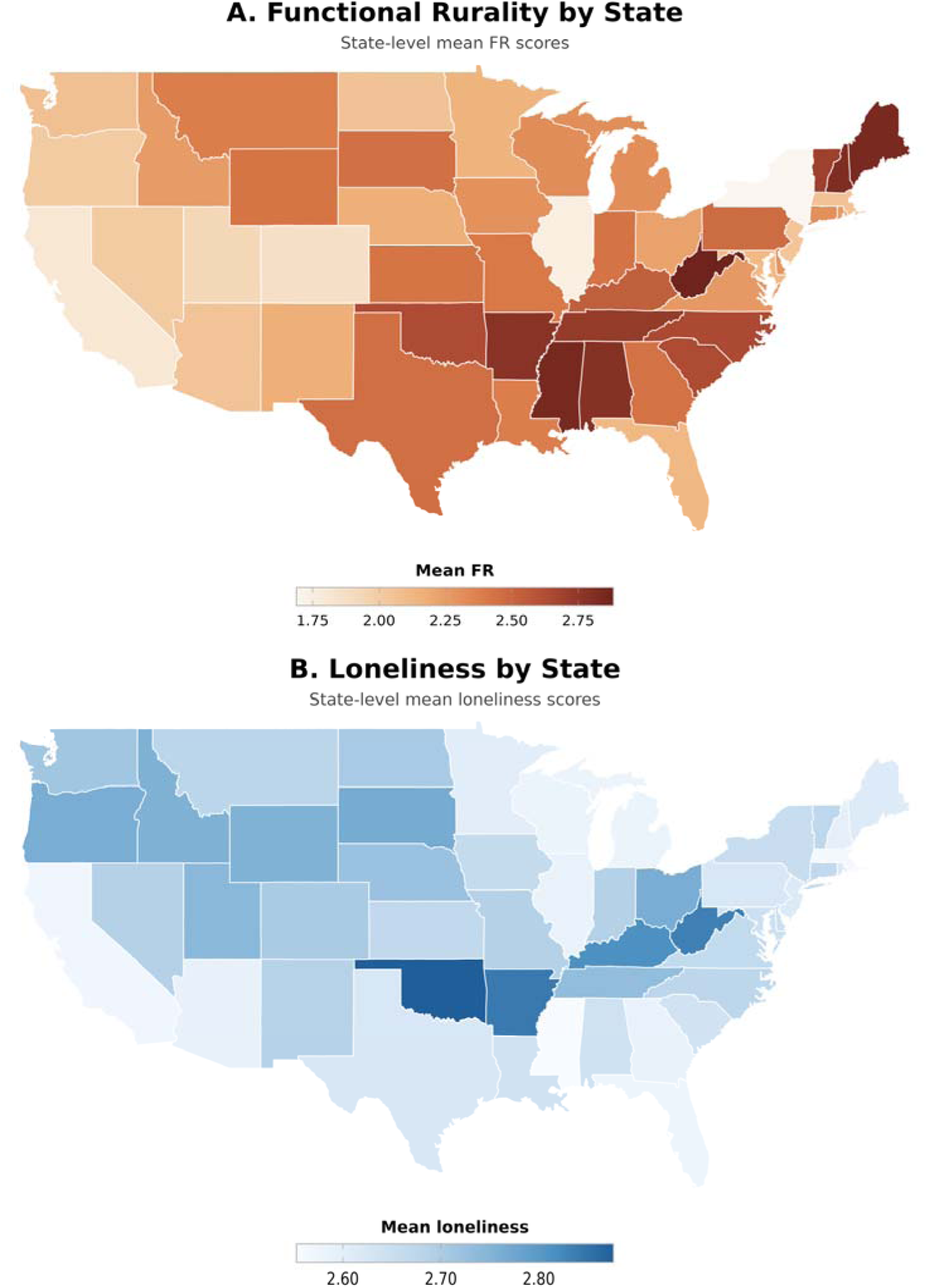
Descriptive state-level maps of mean FR and mean loneliness in the analytic sample.

### Sensitivity Analyses

Sensitivity analyses were directionally consistent with the primary findings. Applying stricter wearable-data requirements (≥10 valid physical activity days or nights) yielded highly similar estimates, as did winsorizing continuous outcomes. Quartile-based exposure models showed broadly similar dose-response patterns, and analyses using a backward 30-day window showed the same overall directions of association with some attenuation in magnitude. These supplementary analyses therefore supported the robustness of the primary findings. Supplementary Result S1 is included in supplementary material.

## Discussion

In a series of preregistered cross-sectional analyses using SDoH survey data linked with overlapping Fitbit data from the All of Us Research Program, we found that higher FR was associated with greater loneliness and that both FR and loneliness were independently associated with lower physical activity. In contrast, loneliness showed the clearest and most consistent associations with sleep: participants with greater loneliness slept fewer hours on average, had greater night-to-night variability in sleep duration, and had higher odds of short sleep and low sleep efficiency. Evidence for effect modification by FR was limited overall, with the notable exception of sleep duration variability, for which higher FR strengthened the positive association between loneliness and irregular sleep duration.

A classic theme in urban sociology is whether denser, more connected, and more accessible environments foster greater social integration by increasing opportunities for incidental contact and the formation of “weak ties,” that is, routine, low-intensity interactions that emerge in everyday settings such as sidewalks, shops, and public transit.^30^ Although brief, these interactions may help sustain a baseline sense of social embeddedness and belonging.^12^ In contrast, environments characterized by reduced accessibility and fewer shared public spaces may limit opportunities for such incidental encounters, thereby constraining the formation and maintenance of weak ties.

From this perspective, FR can be understood as an accessibility-based constraint on the social ecology of daily life. The FR construct used here captures features of the built environment, including limited walkability, reduced transit access, and fewer nearby destinations, that may diminish the frequency of casual social exposure. This is one plausible explanation for the positive association between FR and loneliness observed in the present study. FR environments may be “lonelygenic” insofar as they systematically reduce opportunities for low-effort social contact, even when individuals are not explicitly socially withdrawn. This framing is consistent with recent work suggesting that features of the built environment may shape loneliness not only through social composition, but also through the structure of everyday interaction opportunities.^11^

Importantly, this interpretation does not imply that rural or low-access environments are inherently socially impoverished. Rather, it suggests that the type and distribution of social contact may differ, with fewer ambient or incidental interactions and greater reliance on intentional, effortful social engagement, including forms of connection that are not captured by FR, such as church involvement, community groups, and other organized social activities. In such contexts, individuals who are already vulnerable to loneliness may be particularly affected because the environmental scaffolding that supports routine social connection is less available. The small but consistent association between higher FR and greater loneliness is therefore compatible with a broader ecological view in which built-environment features shape the baseline probability of everyday social contact and, by extension, perceived social connection.

Sleep outcomes were more strongly patterned around loneliness than around FR. In both separate and mutually adjusted models, loneliness showed consistent associations with shorter sleep duration, greater sleep duration variability, and higher odds of short sleep and low sleep efficiency, whereas FR showed minimal relationships with most sleep parameters. This pattern suggests that sleep, as a behavioral and physiological outcome, may be more tightly coupled to psychosocial experience than to the accessibility features captured by FR.

This interpretation is consistent with theoretical accounts that conceptualize loneliness as a state of heightened social vigilance. From this perspective, loneliness reflects not simply a deficit in social contact, but a perceived threat to social safety that activates sustained monitoring of the environment. Such vigilance may interfere with the downregulation of arousal necessary for restorative sleep, contributing to shorter, less regular, and less efficient sleep.^1^ Meta-analytic evidence likewise indicates that loneliness is reliably associated with poorer subjective and objective sleep, including shorter duration and greater disturbance.^5^ Prior work has linked loneliness and social isolation to objectively measured sleep disturbance, but much of that evidence has come from smaller or more selected samples. The present findings place this association on stronger empirical footing by showing that loneliness is consistently related to multiple wearable-derived sleep indicators in a large and diverse U.S. cohort under free-living conditions. In this sense, our results do not merely replicate prior work; they strengthen the evidence that loneliness is robustly associated with adverse sleep patterns in daily life, including night-to-night variability in sleep duration, which may reflect instability in daily routines or circadian regulation.

The relative absence of associations between FR and most sleep outcomes further underscores the distinction between environmental constraint and psychosocial mechanism. Whereas features of the built environment may shape opportunities for movement and incidental social contact, their influence on sleep may be more indirect and contingent. Sleep occurs in a more bounded and private context, and may therefore be less sensitive to neighborhood accessibility per se than to internal states such as perceived safety, stress, and emotional regulation. To the extent that FR does influence sleep, it may do so through unmeasured pathways, such as work schedules, commuting demands, environmental noise, or light exposure, that were not directly captured in the present analysis. The small positive association between FR and sleep efficiency should therefore be interpreted cautiously. Its magnitude was minimal, it did not extend to sleep duration or variability, and it may reflect residual selection, measurement constraints in bring-your-own-device data, or unobserved aspects of daily structure rather than a substantively beneficial effect of lower-access environments on sleep. Taken together, these findings point to loneliness as the more robust and proximal correlate of sleep disruption, consistent with models that locate sleep at the intersection of social experience and physiological regulation.

The physical activity findings suggest that contextual accessibility and psychosocial experience may each contribute to lower movement through partially distinct pathways. FR, as defined here, reflects reduced access to walkable destinations, transit, sidewalks, bicycling facilities, and low-cost recreation, which are features commonly implicated in activity-supportive environments.^10^ Loneliness may operate more through behavioral withdrawal, reduced social motivation, or fewer opportunities for shared activity. The persistence of both FR and loneliness in mutually adjusted models is therefore consistent with the idea that the built environment and social experience each matter for everyday physical activity.

The interaction analyses suggest that FR generally did not amplify the behavioral correlates of loneliness in this sample. The one exception, sleep duration variability, indicates that the positive association between loneliness and irregular sleep duration became stronger at higher levels of FR. This pattern suggests that lower-access environments may provide fewer opportunities for routine incidental connection that could otherwise buffer the sleep-related consequences of loneliness. At the same time, because most other interaction terms were null, this finding should be interpreted conservatively until replicated.

This study has several strengths. It used a large wearable-linked cohort, synchronized exposure measurement from the same SDoH survey instance, objective free-living activity and sleep summaries, and multiple prespecified sensitivity analyses. In addition, to our knowledge, this study is among the first to leverage the All of Us Research Program to operationalize an FR construct in a large and diverse U.S. cohort. This approach extends prior work on neighborhood accessibility and rural context by providing an initial U.S.-based implementation of FR using built-environment and access-related SDoH items. At the same time, several limitations should be noted. First, the design is cross-sectional, so temporal ordering and causality cannot be established. Second, the analytic sample represents a selected subset of All of Us participants who completed the optional SDoH survey, linked a personal Fitbit, and met wear criteria; results should therefore not be interpreted as population-representative estimates. Third, FR is a study-specific contextual index rather than a validated geographic rurality classification and should be interpreted as a proxy for reduced neighborhood accessibility. Fourth, Fitbit metrics are useful but imperfect behavioral proxies, especially for sleep parameters.^21,27,28^ Reverse causation and selection processes are also possible. For example, individuals who are dispositionally more socially withdrawn or loneliness-prone may be more likely to sort into lower-density or lower-access environments, which could contribute to the observed associations. Additionally, FR does not capture all forms of social engagement, such as religious participation, community organizations, or other intentional forms of connection that may remain important in lower-access settings. Finally, complete-case analysis may have introduced additional selection biases.

Despite these limitations, the findings have practical implications. Efforts to improve physical activity may benefit from attending to both neighborhood accessibility and psychosocial isolation, whereas efforts to improve sleep may need to more directly address loneliness and related social stressors. More broadly, the study demonstrates the value of linking social determinants data with wearable measures in large, diverse cohorts to examine everyday behavioral pathways relevant to chronic disease prevention.

### Conclusion

Higher functional rurality and higher loneliness were each associated with lower Fitbit-measured physical activity in this All of Us sample, whereas loneliness showed the more consistent and unfavorable pattern of association across sleep outcomes. Functional rurality was associated with greater loneliness but showed only limited and mixed associations with sleep. The positive association between loneliness and sleep duration variability was stronger in higher-FR contexts, although most interaction terms were null. Taken together, these findings highlight the importance of considering both contextual accessibility and psychosocial experience when examining health-relevant behavioral correlates.

## Data Availability

The data analyzed in this study are not publicly available because they are participant-level human data from the All of Us Research Program Controlled Tier Dataset. Eligible researchers may obtain access through the All of Us Researcher Workbench upon application and approval. Summary results are provided in the manuscript and supplementary materials.

## Author Contributions

Shuai Yang had full access to all the study data and takes full responsibility for the integrity and accuracy of the reporting.

Conceptualization: Shuai Yang (SY), David A. Sbarra (DAS), Yann C. Klimentidis (YCK)

Methodology: SY, DAS, YCK

Data curation: SY

Software: SY

Formal analysis: SY

Visualization: SY

Writing – original draft: SY

Writing – review & editing: All authors

Supervision: DAS, YCK

## Transparncy and Openness Statement

This study was preregistered at OSF: https://osf.io/9857p/overview?view_only=477e2e2f76c340fe8aadbee284064f07. Because the study used restricted-access data from the All of Us Research Program, participant-level data are not publicly available. Analytic code and variable-construction scripts are available at [https://osf.io/9857p/files/osfstorage/69d5b0f1e04108719033eb1b]. Study materials, including variable definitions and supplementary analytic details, are provided in the manuscript and Supplementary Materials. Deviations from the preregistered plan are described in the Supplementary Materials.

## Generative AI Use Statement

No generative AI tools were used in the drafting, analysis, or preparation of this manuscript.

