## Supplementary Material for "Loneliness, Functional Rurality, and Wearable-Measured Physical Activity and Sleep in the All of Us Research Program"

**Supplementary** **Table S1. Associations of functional rurality and loneliness with physical activity and sleep outcomes**

| **Outcome** | **Model** | **Exposure** | **Estimate** | **95% CI** | ***p*** |
| --- | --- | --- | --- | --- | --- |
| Daily steps | Separate | FR | β = -211.23 | [-268.75, -153.72] | 6.33 × 10^-13^ |
| Daily steps | Separate | Loneliness | β = -380.06 | [-439.78, -320.35] | 1.47 × 10^-35^ |
| Daily steps | Joint | FR | β = -187.49 | [-244.73, -130.24] | 1.41 × 10^-10^ |
| Daily steps | Joint | Loneliness | β = -366.64 | [-426.27, -307.02] | 2.56 × 10^-33^ |
| MVPA minutes | Separate | FR | β = -1.78 | [-2.30, -1.25] | 3.96 × 10^-11^ |
| MVPA minutes | Separate | Loneliness | β = -1.66 | [-2.19, -1.13] | 6.19 × 10^-10^ |
| MVPA minutes | Joint | FR | β = -1.68 | [-2.20, -1.15] | 3.84 × 10^-10^ |
| MVPA minutes | Joint | Loneliness | β = -1.54 | [-2.06, -1.02] | 7.90 × 10^-9^ |
| Sleep duration (hours) | Separate | FR | β = 0.003 | [-0.016, 0.021] | .766 |
| Sleep duration (hours) | Separate | Loneliness | β = -0.093 | [-0.113, -0.073] | 4.40 × 10^-19^ |
| Sleep duration (hours) | Joint | FR | β = 0.009 | [-0.010, 0.027] | .363 |
| Sleep duration (hours) | Joint | Loneliness | β = -0.094 | [-0.114, -0.073] | 2.95 × 10^-19^ |
| Sleep efficiency | Separate | FR | β = 0.00078 | [0.00024, 0.00132] | .0045 |
| Sleep efficiency | Separate | Loneliness | β = -0.00047 | [-0.00104, 0.00011] | .114 |
| Sleep efficiency | Joint | FR | β = 0.00081 | [0.00027, 0.00135] | .0032 |
| Sleep efficiency | Joint | Loneliness | β = -0.00052 | [-0.00110, 0.00006] | .0778 |
| Sleep duration variability (hours) | Separate | FR | β = 0.0038 | [-0.0066, 0.0143] | .472 |
| Sleep duration variability (hours) | Separate | Loneliness | β = 0.0704 | [0.0592, 0.0817] | 2.90 × 10^-34^ |
| Sleep duration variability (hours) | Joint | FR | β = -0.0005 | [-0.0109, 0.0099] | .928 |
| Sleep duration variability (hours) | Joint | Loneliness | β = 0.0705 | [0.0592, 0.0818] | 3.43 × 10^-34^ |
| Short sleep | Separate | FR | OR = 0.998 | [0.962, 1.035] | .904 |
| Short sleep | Separate | Loneliness | OR = 1.141 | [1.097, 1.186] | 3.08 × 10^-11^ |
| Short sleep | Joint | FR | OR = 0.989 | [0.954, 1.026] | .568 |
| Short sleep | Joint | Loneliness | OR = 1.142 | [1.098, 1.187] | 2.72 × 10^-11^ |
| Low sleep efficiency | Separate | FR | OR = 0.934 | [0.882, 0.988] | .0172 |
| Low sleep efficiency | Separate | Loneliness | OR = 1.107 | [1.045, 1.173] | 5.95 × 10^-4^ |
| Low sleep efficiency | Joint | FR | OR = 0.928 | [0.877, 0.982] | .0099 |
| Low sleep efficiency | Joint | Loneliness | OR = 1.112 | [1.049, 1.179] | 3.46 × 10^-4^ |

***Note****. Continuous exposures were standardized (1-SD increase). Linear models are reported as β coefficients; logistic models are reported as odds ratios (ORs). All models adjusted for age, gender, race, education, income, smoking, alcohol use, and season of t0. Robust HC3 standard errors were used for inference.*

**Supplementary** **Table S2. State-level mean functional rurality and loneliness in the analytic sample**

| **State** | **N** | **Mean FR** | **Mean loneliness** |
| --- | --- | --- | --- |
| Alabama | 9,215 | 2.795 | 2.640 |
| Alaska | 182 | 2.372 | 2.762 |
| Arizona | 16,350 | 2.051 | 2.588 |
| Arkansas | 604 | 2.780 | 2.842 |
| California | 31,669 | 1.813 | 2.570 |
| Colorado | 4,230 | 1.864 | 2.702 |
| Connecticut | 1,526 | 2.306 | 2.666 |
| Delaware | 364 | 2.292 | 2.651 |
| Florida | 9,305 | 2.122 | 2.576 |
| Georgia | 5,934 | 2.439 | 2.586 |
| Hawaii | 462 | 2.145 | 2.623 |
| Idaho | 455 | 2.252 | 2.755 |
| Illinois | 20,587 | 1.757 | 2.585 |
| Indiana | 2,708 | 2.433 | 2.689 |
| Iowa | 1,432 | 2.300 | 2.662 |
| Kansas | 1,813 | 2.424 | 2.665 |
| Kentucky | 1,205 | 2.538 | 2.813 |
| Louisiana | 2,788 | 2.387 | 2.639 |
| Maine | 761 | 2.843 | 2.613 |
| Maryland | 2,982 | 2.115 | 2.646 |
| Massachusetts | 17,780 | 2.057 | 2.564 |
| Michigan | 13,814 | 2.313 | 2.579 |
| Minnesota | 6,436 | 2.145 | 2.605 |
| Mississippi | 1,453 | 2.846 | 2.553 |
| Missouri | 1,639 | 2.404 | 2.688 |
| Montana | 355 | 2.384 | 2.679 |
| Nebraska | 590 | 2.166 | 2.722 |
| Nevada | 1,313 | 2.007 | 2.689 |
| New Hampshire | 1,034 | 2.817 | 2.594 |
| New Jersey | 1,306 | 2.040 | 2.616 |
| New Mexico | 1,537 | 2.166 | 2.687 |
| New York | 9,589 | 1.691 | 2.656 |
| North Carolina | 4,062 | 2.651 | 2.676 |
| North Dakota | 434 | 2.057 | 2.704 |
| Ohio | 3,458 | 2.229 | 2.762 |
| Oklahoma | 908 | 2.637 | 2.876 |
| Oregon | 1,870 | 2.007 | 2.762 |
| Pennsylvania | 22,005 | 2.473 | 2.623 |
| Rhode Island | 691 | 2.257 | 2.618 |
| South Carolina | 1,469 | 2.637 | 2.636 |
| South Dakota | 218 | 2.448 | 2.766 |
| Tennessee | 3,509 | 2.734 | 2.732 |
| Texas | 8,566 | 2.454 | 2.623 |
| Utah | 769 | 1.933 | 2.741 |
| Vermont | 270 | 2.703 | 2.672 |
| Virginia | 2,020 | 2.265 | 2.665 |
| Washington | 4,003 | 2.070 | 2.713 |
| West Virginia | 487 | 2.881 | 2.835 |
| Wisconsin | 20,053 | 2.318 | 2.582 |
| Wyoming | 161 | 2.424 | 2.755 |

*Note:* *Values are state-level mean functional rurality (FR) and mean loneliness scores in the analytic sample. Results are presented for descriptive purposes.*

**Supplementary Figure S1. Heatmap of correlations among primary continuous study variables.** Cells display Pearson correlation coefficients for functional rurality (FR), loneliness, mean daily steps, mean daily moderate-to-vigorous physical activity (MVPA) minutes, mean sleep duration, mean sleep efficiency, and sleep duration variability. Color intensity reflects the direction and magnitude of the correlation, and numeric values are shown within each cell.

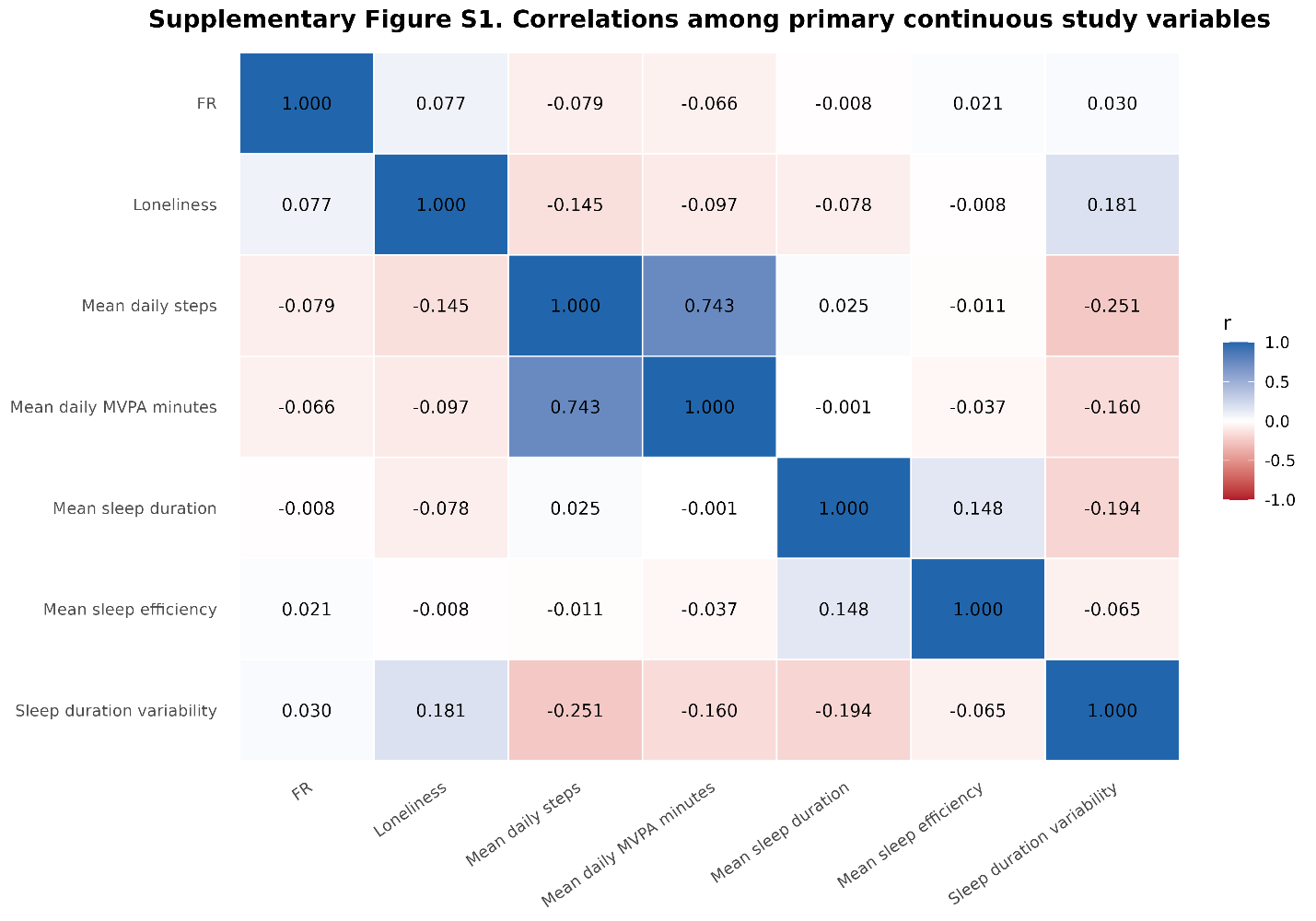

**Supplementary Method 1. Deviations from the Preregistered Analysis Plan**

Below we describe all deviations from the preregistered plan and the rationale for each change.

**1. Addition of gender as a covariate.**
The preregistered analysis plan did not include gender as a covariate. In the final analyses, we added gender to the adjusted models because gender is associated with loneliness, physical activity, and sleep, and therefore represented a plausible confounder of the associations under study. Including gender was intended to improve covariate control and provide more conservative estimates of the associations of functional rurality and loneliness with wearable-derived behavioral outcomes. This change is reported here for transparency.

**Supplementary Result S1: Sensitivity Analyses**

Several sensitivity analyses were conducted to evaluate the robustness of the primary findings. First, applying stricter wearable data requirements (≥10 valid physical activity days or ≥10 valid sleep nights) produced results that were highly consistent with the primary analyses. For physical activity, higher functional rurality remained associated with fewer daily steps (β = -218.57, 95% CI [-277.43, -159.71]) and lower MVPA minutes (β = -2.11, 95% CI [-2.66, -1.56]), while greater loneliness remained associated with fewer daily steps (β = -397.88, 95% CI [-459.98, -335.78]) and lower MVPA minutes (β = -1.77, 95% CI [-2.33, -1.21]). For sleep outcomes, loneliness remained associated with shorter sleep duration (β = -0.086, 95% CI [-0.107, -0.064]), greater sleep duration variability (β = 0.071, 95% CI [0.059, 0.082]), higher odds of short sleep (OR = 1.12, 95% CI [1.08, 1.17]), and higher odds of low sleep efficiency (OR = 1.11, 95% CI [1.05, 1.18]). Functional rurality remained positively associated with sleep efficiency (β = 0.00085, 95% CI [0.00027, 0.00143]) and inversely associated with low sleep efficiency (OR = 0.92, 95% CI [0.87, 0.98]), but remained unrelated to mean sleep duration, sleep duration variability, and short sleep.

Second, results were materially unchanged after winsorizing continuous outcomes rather than relying on the primary exclusion rule for implausible values. In the winsorized analyses, functional rurality remained inversely associated with daily steps (β = -224.13, 95% CI [-278.82, -169.43]) and MVPA minutes (β = -2.13, 95% CI [-2.62, -1.64]), and loneliness remained inversely associated with daily steps (β = -394.85, 95% CI [-451.69, -338.00]), MVPA minutes (β = -1.72, 95% CI [-2.23, -1.22]), and sleep duration (β = -0.092, 95% CI [-0.113, -0.071]), while remaining positively associated with sleep duration variability (β = 0.068, 95% CI [0.057, 0.078]). Functional rurality also remained positively associated with sleep efficiency (β = 0.00076, 95% CI [0.00028, 0.00123]) and inversely associated with low sleep efficiency (OR = 0.93, 95% CI [0.88, 0.98]). Loneliness remained associated with higher odds of short sleep (OR = 1.13, 95% CI [1.09, 1.18]) and low sleep efficiency (OR = 1.11, 95% CI [1.05, 1.17]).

Third, analyses using quartile-based exposure coding yielded a similar overall pattern and suggested dose-response relationships for several outcomes. Compared with the lowest FR quartile, participants in higher FR quartiles had fewer daily steps and lower MVPA, with the strongest contrast observed for MVPA in the highest quartile (β = -6.11, 95% CI [-7.61, -4.62]). Higher FR quartiles were also associated with modestly higher sleep efficiency, and the highest FR quartile was associated with lower odds of low sleep efficiency (OR = 0.81, 95% CI [0.69, 0.94]). For loneliness, higher quartiles were associated with progressively fewer daily steps and lower MVPA, particularly in the highest quartile. The highest loneliness quartile was also associated with shorter sleep duration (β = -0.229, 95% CI [-0.287, -0.171]), greater sleep duration variability (β = 0.170, 95% CI [0.139, 0.200]), higher odds of short sleep (OR = 1.40, 95% CI [1.26, 1.56]), and higher odds of low sleep efficiency (OR = 1.31, 95% CI [1.12, 1.54]).

Finally, using a backward 30-day window before t0 also produced directionally similar findings, although effect sizes were somewhat attenuated because of the smaller analytic sample. In these analyses, both functional rurality and loneliness remained inversely associated with daily steps and MVPA. Loneliness remained associated with shorter sleep duration (β = -0.071, 95% CI [-0.096, -0.045]), greater sleep duration variability (β = 0.064, 95% CI [0.051, 0.077]), and higher odds of short sleep (OR = 1.07, 95% CI [1.02, 1.12]) and low sleep efficiency (OR = 1.09, 95% CI [1.02, 1.17]). Functional rurality was again not meaningfully associated with mean sleep duration or sleep duration variability, and its association with low sleep efficiency was attenuated and no longer statistically significant in the backward-window analysis (OR = 0.96, 95% CI [0.90, 1.03]). Overall, these sensitivity analyses support the robustness of the primary findings, particularly for the inverse associations of both functional rurality and loneliness with physical activity and for the associations of loneliness with shorter and more variable sleep.
